# The unique evolutionary dynamics of the SARS-CoV-2 Delta variant

**DOI:** 10.1101/2021.08.05.21261642

**Authors:** Adi Stern, Shay Fleishon, Talia Kustin, Edo Dotan, Michal Mandelboim, Oran Erster, Israel Consortium of SARS-CoV-2 sequencing, Ella Mendelson, Orna Mor, Neta S. Zuckerman

**Affiliations:** The Shmunis School of Biomedicine and Cancer Research, George S. Wise Faculty of Life Sciences, Tel Aviv University, Tel Aviv, Israel; Edmond J. Safra Center for Bioinformatics, Tel Aviv University, Tel Aviv, Israel; Central Virology Laboratory, Public Health Services, Ministry of Health and Sheba Medical Center, Tel-Hashomer, Israel; School of Public Health, Sackler Faculty of Medicine, Tel-Aviv University, Tel-Aviv, Israel

**Author notes:** Co-equal authorship (Adi Stern, Shay Fleishon). Israel Consortium of SARS-CoV-2 sequencing: Neta S. Zuckerman, Orna Mor, Efrat Dahan Bucris, Michal Mandelboim, Danit Sofer, Dana Bar-Ilan, Miranda Geva, Omer Asraf, Oran Erster, Gideon Rechavi, Efrat Glick-Saar, Nir Rainy, Chen Weiner, Reut Sorek-Abramovich, Yevgeni Yegorov, Anna Vishnevsky, Patricia Benveniste-Lekovitz, Abu Hamad Ramzia, Adina Bar Chaim, Ella Mendelson.

## Abstract

The SARS-Coronavirus-2 (SARS-CoV-2) driven pandemic was first recognized in late 2019, and the first few months of its evolution were relatively clock-like, dominated mostly by neutral substitutions. In contrast, the second year of the pandemic was punctuated by the emergence of several variants that bore evidence of dramatic evolution. Here, we compare and contrast evolutionary patterns of various variants, with a focus on the recent Delta variant. Most variants are characterized by long branches leading to their emergence, with an excess of non-synonymous substitutions occurring particularly in the Spike and Nucleocapsid proteins. In contrast, the Delta variant that is now becoming globally dominant, lacks the signature long branch, and is characterized by a step-wise evolutionary process that is ongoing. Contrary to the “star-like” topologies of other variants, we note the formation of several distinct clades within Delta that we denote as clades A-E. We find that sequences from the Delta D clade are dramatically increasing in frequency across different regions of the globe. Delta D is characterized by an excess of non-synonymous mutations, mostly occurring in ORF1a/b, some of which occurred in parallel in other notable variants. We conclude that the Delta surge these days is composed almost exclusively of Delta D, and discuss whether selection or random genetic drift has driven the emergence of Delta D.

## Introduction

The COVID-19 pandemic was first recognized in Wuhan, China in December 2019 and the etiological agent of the disease SARS-Coronavirus-2 was sequenced around ten days after the disease was formally discovered [1]. During the few months of the pandemic the evolution of the virus was relatively predictable, with substitutions accumulating at a fixed pace of about one substitution every second week [2][3][4][5], at a rate compatible with a molecular clock based on neutral evolution, and no evidence of dramatic positive selection was observed [6]. One notable exception was the D614G substitution in the Spike (S) protein, which quickly rose to fixation and indeed later evidence showed that this mutation is associated with increased transmissibility [7][8][9][10]. However, since the fall of 2020, several SARS-CoV-2 variants have emerged with particular genomic and epidemiological features, all suggestive of some form of selection operating on them. The first such variant was B.1.1.7 (Alpha), first detected in the U.K. Its most prominent characteristic was the overall wealth of substitutions found in it, especially in the region encoding for S [11]. Subsequently, it was found that B.1.1.7 ubsequently, it was found that B.1.1.7 spreads exceptionally rapidly [12][13] and its high transmissibility led to it displacing the originally circulating strains in many different countries around the world.

Overall, variants are characterized based on well-defined phylogenetic clades, and classified based on available evidence for increased transmissibility, virulence, or escape from the immune system / therapeutics. Variants with these characteristics are classified as variants of concern (VOC), variants of interest (VOI) or variants under monitoring (VUM) by health agencies such as the world health organization (WHO, www.who.int). Thus far, the WHO has defined four VOCs: B.1.1.7 (Alpha), B.1.351 (Beta), P.1 (Gamma), B.1.617.2 (Delta), and four VOIs: B.1.525 (Eta), B.1.526 (Iota), B.1.617.1 (Kappa) and C.37 (Lambda). An additional twelve variants previously classified as VOIs, such as B.1.427/9 originating in California [14], have now been reclassified as VUMs. For the purpose of our investigation herein, we collectively denote a VOC/VOI/VUM as a VO.

Following the detection of B.1.1.7, there has been a surge of different variants reported globally, which were characterized by punctual rises in frequencies of a particular variant, often accompanied by its demise a few months later. In the past few months, focus has turned to the Delta variant, which was first detected in India and has recently increased in prevalence globally. The Delta variant currently seems to be displacing all other variants, including the highly dominant and contagious Alpha variant, in numerous countries across the globe [15]. In this study, we utilize evolutionary genomics to explain the rise and fall of different variants across time, with an emphasis on exploring the patterns of evolution in the globally-ascending Delta variant in comparison to other VOs.

## Results

### Genomic features common and unique across VOs

We begin with a global analysis of a representative sample of SARS-CoV-2 sequences. Within the phylogeny of these sequences, most tips (Fig. 1; white dots) are spread across the entire time-line of the tree, with short branches separating the various clades. This reflects the intense sampling of isolates, coupled with the molecular clock of the virus, which dictates, on average, approximately one substitution every second week [2]. However, the branches leading to the VOs represented in the tree are exceptionally long, representing the accumulation of many substitutions. We find that these branches share the following common features: (1) most have a higher proportion of non-synonymous (NS) substitutions as compared to non-VOs (Fig. 2A) (p=0.01, t-test (Methods)), (2) the entire 3’ portion of the genome is enriched for NS substitutions, with a particular emphasis on the S and N genes, but also on ORF3a and ORF8 (Fig. 2B), (3) NS substitutions in S are mostly located in the N terminal domain (NTD) and receptor binding domain (RBD), (Fig. 2C), and (4) often, parallel substitutions are observed among the various VOs, such as at positions 452, 484, or 501 of the Spike protein [16].

**Figure 1.**
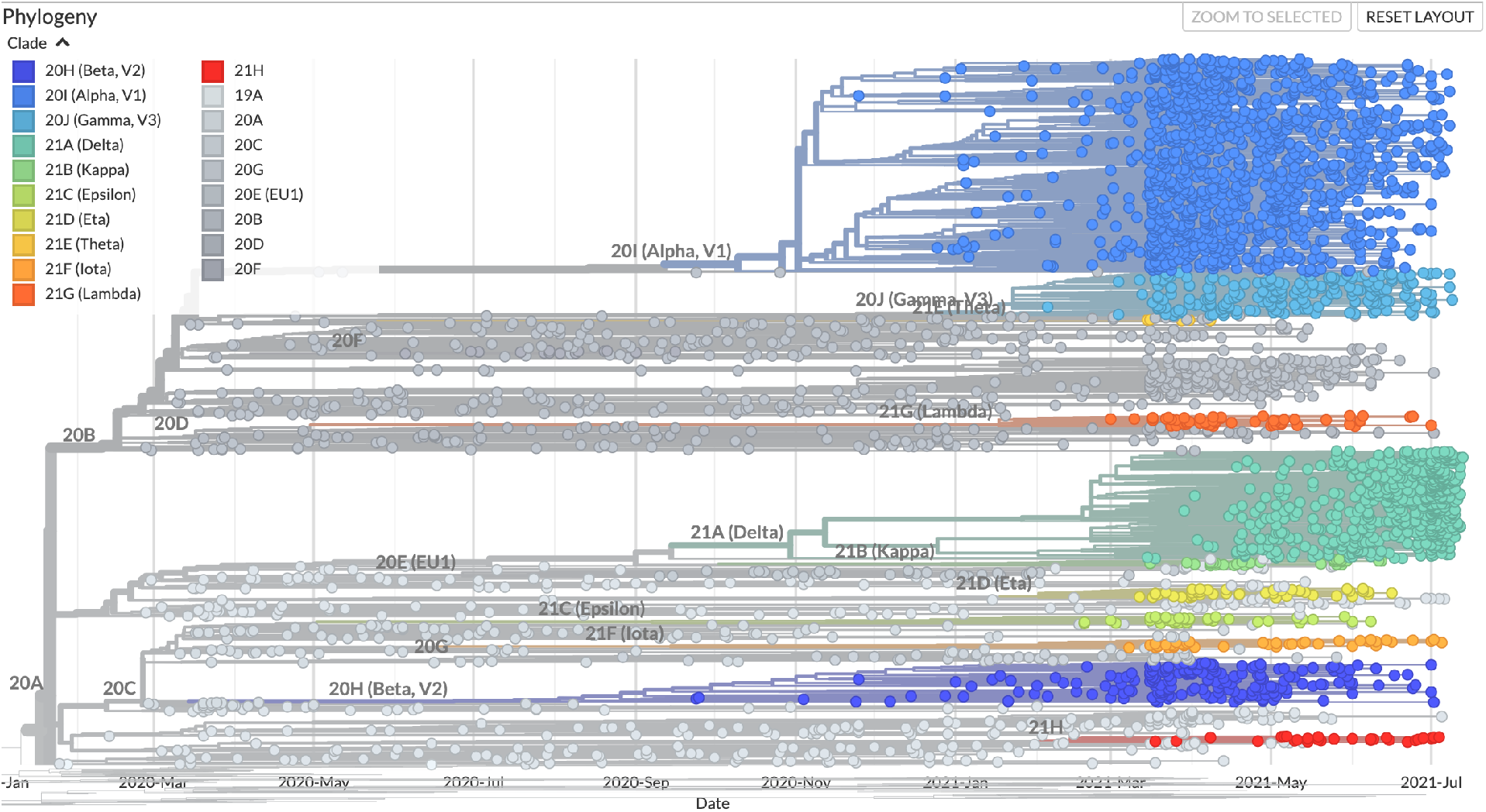
Time-aligned phylogeny of a representative sample of SARS-CoV-2 isolates. Different clades corresponding to VOCs and VOIs are colored. Notably, long branches lead to most VOCs/VOIs, suggesting an increased rate of evolution leading to their emergence. The phylogeny was generated using Nextstrain [18] on July 24 2021.

**Figure 2.**
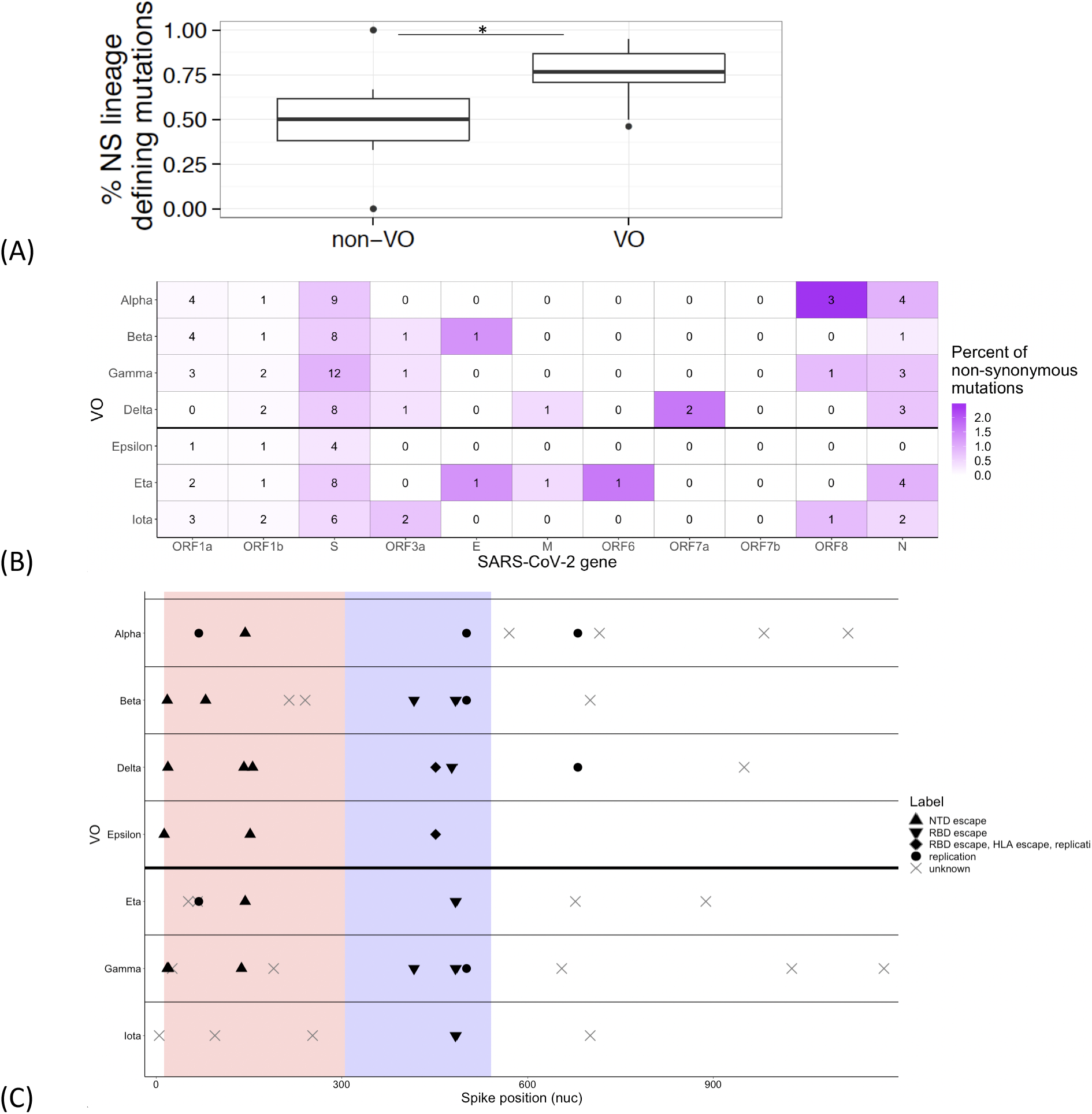
Comparison of lineage defining non-synonymous (NS) substitutions across VOs. (A) The proportion of NS (out of synonymous and non-synonymous substitutions) in VOs compared to non-VO B.1 lineages (Methods) (B) Summary of the number of non-synonymous substitutions per gene. Each cell is colored by the relative proportion of non-synonymous substitutions normalized by protein length. Deletions affecting coding regions are counted as non-synonymous in (A) and (B). (C) Non-synonymous substitutions with a focus on the spike protein. Pink and violet shading correspond to the NTD and RBD, respectively. Each substitution is shaped based on current evidence for the functional change it entails (Table S1), focusing on association with two global categories: escape from antibodies (squares and circles) and enhanced replication (regardless of the mechanisms). The D614G amino-acid substitution shared by all VOs was omitted for clarity.

When focusing on the Spike protein, we found that all VOs are characterized by substitutions prevalent particularly in NTD and (RBD), both critical targets of neutralizing antibodies [17] (Fig. 2C). Interestingly, the one exception is the Alpha VOC, which bears only one mutation associated with immune evasion (a deletion at position 144).

All VOs bear an amino-acid replacement at either position 203, 204, or 205 of the nucleocapsid protein (N), often combined. While it has previously been suggested that the K203/R204 polymorphisms create a non-canonical sgRNA [19], this is not expected to be the case for either the Delta or the Beta variants (Fig. S1). We thus suggest that the amino-acid replacements themselves may be adaptive. Accumulating evidence suggest that N has a crucial role in evasion from the cell autonomous innate immune response [20], [21], in viral assembly [22], and in interactions with cellular co-factors [23], and we suggest that amino-acid replacements at this region may increase replication capacity of the virus.

### Focus on the Delta variant and contrast with Alpha

Both the Delta and Alpha variants (corresponding to NextStrain lineages 21A and 20I, respectively), which spread worldwide, were inferred to have arisen first around the fall of 2020 (with confidence intervals spanning spring 2020 through winter 2020/2021) [18]. However, when analyzing the phylogenies of each variant, the tree topologies looks vastly different (Fig. 3A). VO phylogenies are characterized by multiple polytomies (“star”-like phylogenies) (Fig. 3A, Fig. S2), whereas the Delta phylogeny is more structured. The signature long branch leading to VOs, as described above, is absent in Delta, and it appears that some of the key signature substitutions associated with Delta (e.g., S: L452R, S: P681R) were created in a series of independent steps, as evidenced by multiple shared substitutions with the Kappa variant and other sequences. In fact, when performing a thorough analysis of all globally available Delta sequences (Methods), we were able to separate the Delta phylogeny into five distinct clades, which we label Delta A through E, each characterized by a specific set of substitutions (Methods) (Fig. 3B, Table 1). These clades encompass the three recently noted VOIs AY.1, AY.2 and AY.3 (Fig. 3A) and all other AY lineages defined by the Pango nomenclature system [24].

**Figure 3.**
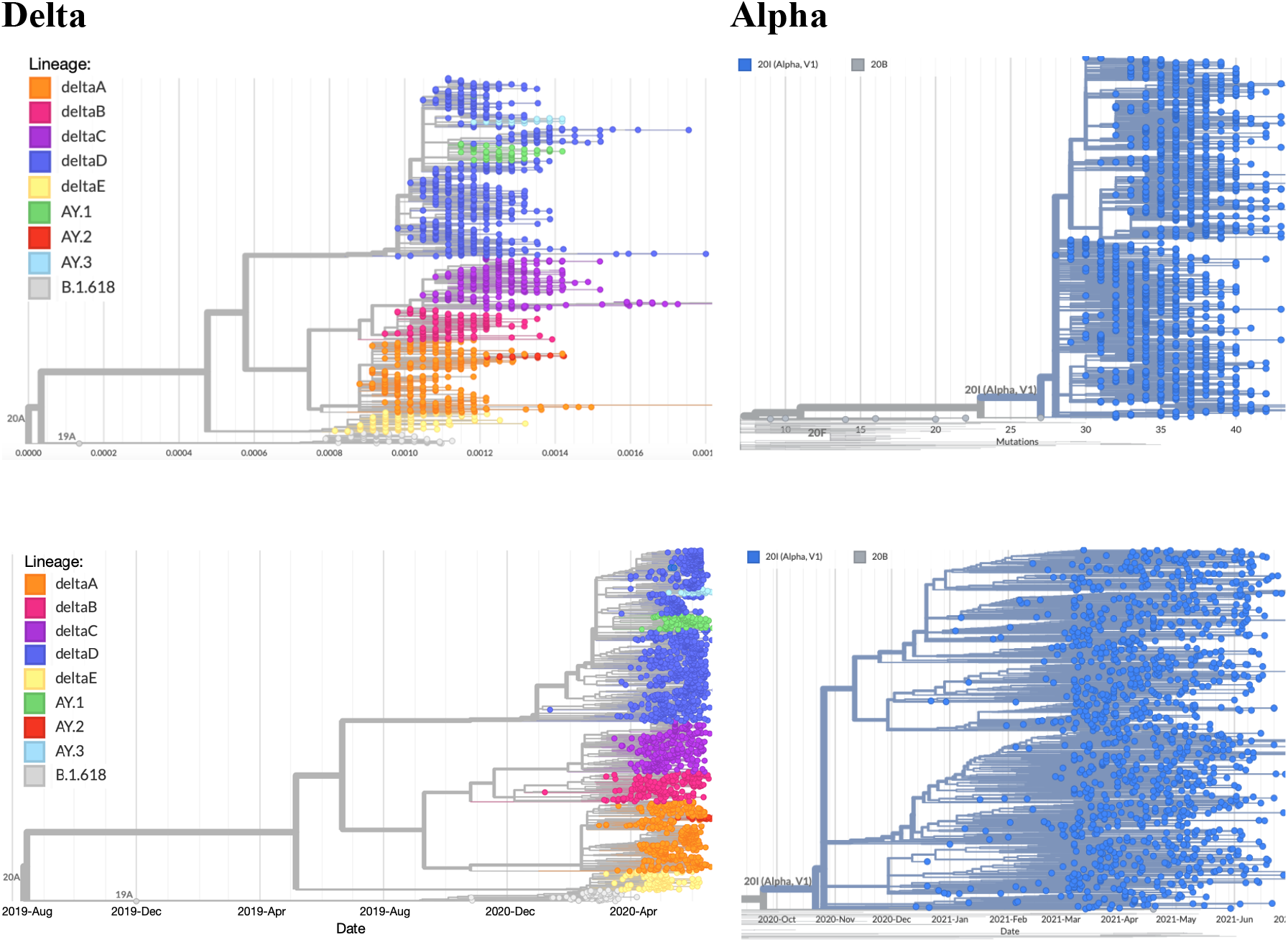
Phylogenies of the Delta and Alpha variants. The two upper panels are divergence-based phylogenies and the lower panels are time-aligned phylogenies. Clades in Delta are separated based on long branches or based on sub-variant designations (see main text) and color-coded accordingly. The B.1.618 was used as an outgroup for the Delta phylogeny, whereas 20B sequences were used as an outgroup for the Delta phylogeny. Substitutions defining the five clades of Delta A-E are specified in Table 1.

**Table 1.**
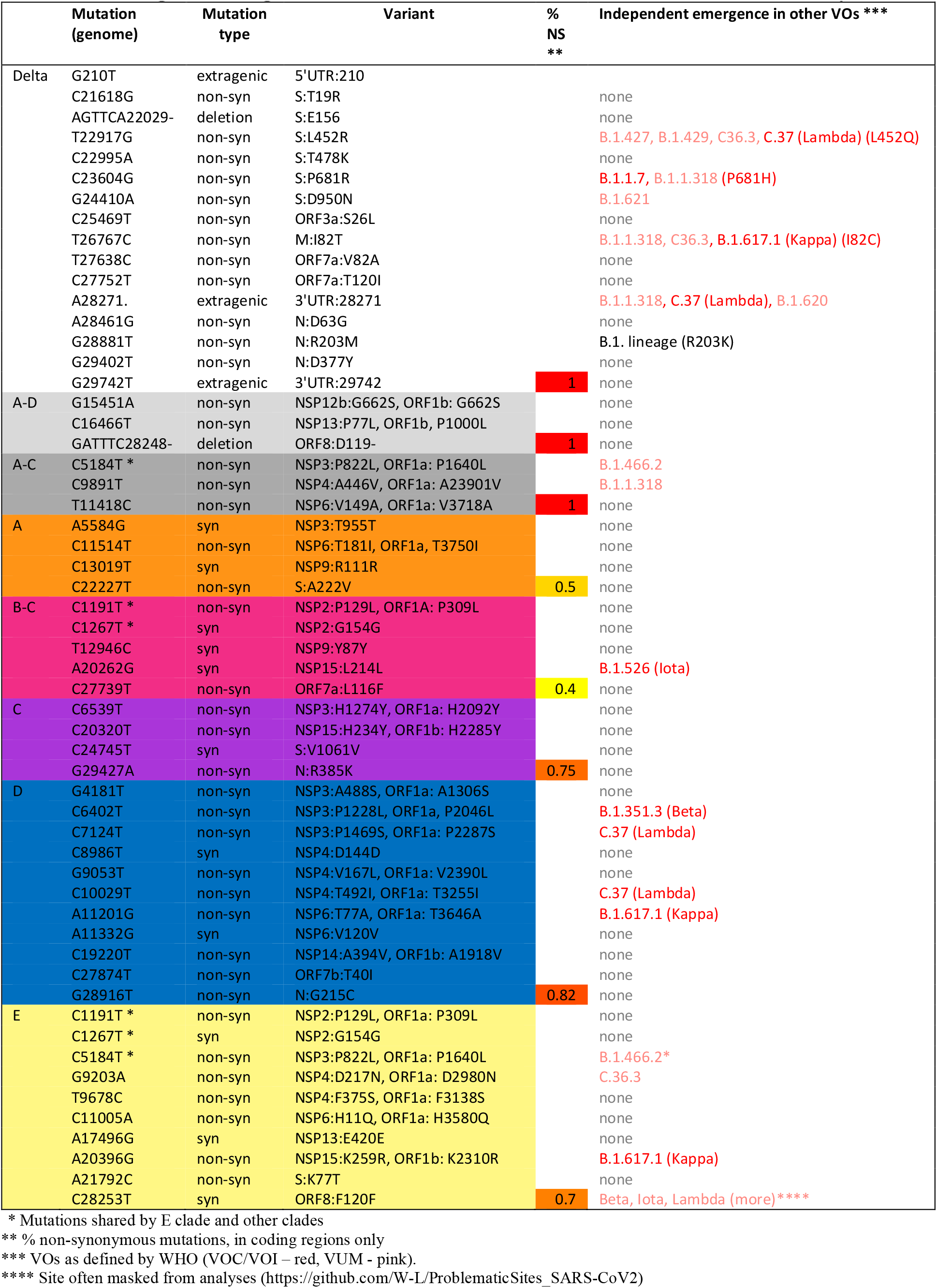
Lineage defining mutations of Delta and its clades defined in this study.

We went on to examine the lineage defining mutations of each clade. We first noted a high proportion of non-synonymous substitutions in some of the clades, in particular clades D and E, which are both characterized by a large number of overall lineage-defining mutations (Table 1). Interestingly, concurrent clade E sequences appear to be a result of recombination between an ancestral predicted E sequence and an ancestor of the B+C clade, since all clade E sequences share the substitutions unique to the B+C clade in the first ∼5200 bases of their genomes (Table 1).

Next, we observed that when focusing on the *basal* Delta lineage (i.e., the branch leading to the emergence of the Delta variant), many lineage defining mutations also occurred independently in other VO lineages, in line with this being one of the four signatures we described above for all VOs. When zooming in to the five Delta clades and their associated lineage defining mutations, we found that theses mutations were also found in other VOs. This was particularly true for Delta D clade which is defined by four mutations common to other VOCs/VOIs (Table 1). This interpretation requires caution, since it does not consider the probability of independent acquisition of mutations due to the high mutation rate of the virus, or other technical artefacts of sequencing.

The prevalence of the five newly characterized Delta clades A-E was analyzed in several countries where the frequency of the Delta variant has been rapidly increasing over time [15] (Fig. S3). All countries exhibited a similar pattern, in which the Delta D clade has gained dominance over the other Delta clades (A,B,C,E) over time (Fig. 4A). In India, where this variant was first discovered, all the Delta clades initially co-occur, with the Delta D clade gaining dominance only in May. We note that the first sequences bearing the Delta D lineage defining mutations are from February-March 2021, and are predominantly from India, suggesting this clade was first created there.

**Figure 4.**
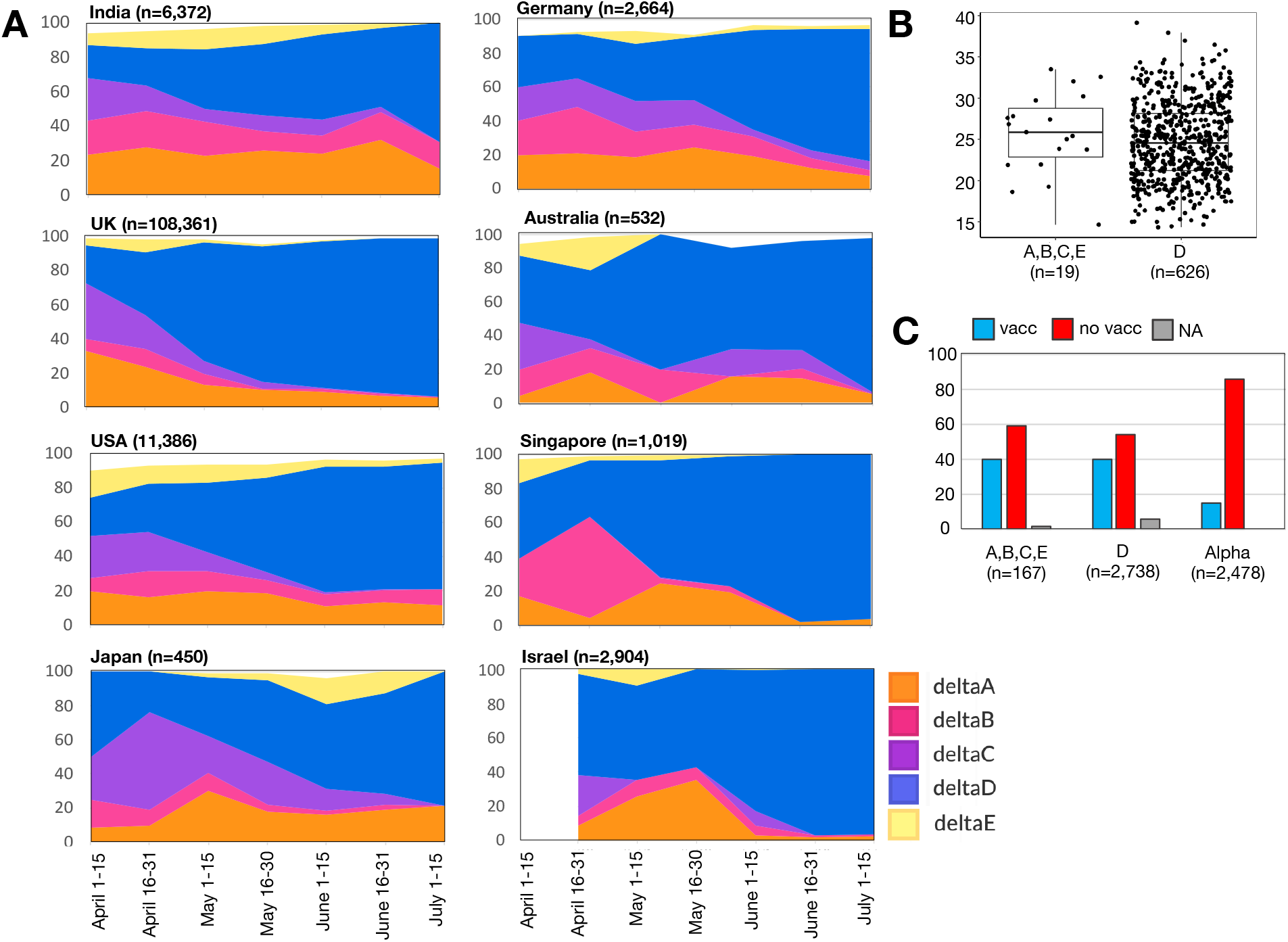
Prevalence and characteristics of Delta clades. (A) Frequency of the five Delta clades A-E between April and July 2020 in countries where the Delta variant has rapidly increased. Frequencies were calculated per time-point, where non-complete/corrupted sequences that could not be classified as any of the Delta clades were removed from the analysis but not from the number of total sequences, hence not all frequencies sum up to 100%. The number of the total sequences evaluated per country is indicated in parentheses. (B) A comparison of cycle threshold (Ct) values from individuals infected by Delta D versus other clades of Delta, (C) breakdown of the proportion of vaccinated versus non- vaccinated individuals infected by Delta D, other clades of Delta and the Alpha variant.

### Interrogation of Delta D infection characteristics

Next, we explored the characteristics of Delta infections in Israel, comparing Delta clade D sequences to other Delta clades. No significant differences were found in viral load, measured by the cycle threshold (Ct), between Delta D infections and infections from the other Delta clades (Fig. 4C), with the caveat that only a small number of samples had available Ct values for the non-clade D infections. Similarly, no striking differences were observed between Delta D and Delta A-E in age or gender (Table S2). We next focused on the proportion of vaccinated (two vaccine doses) and non-vaccinated individuals infected with the Delta variant. When comparing overall Delta clades with Alpha, it was notable that a higher proportion of vaccinated individuals become infected with Delta as compared to Alpha, whereas no notable differences were observed between Delta D and other Delta clades (Fig. 4C, Table S2). These results are line with reports on lower vaccine effectiveness against infection with regards to the Delta variant [25][26][27], but do not add any information why Delta D is more prevalent.

## Discussion

The identification of new SARS-CoV-2 variants is justifiably causing constant trepidation around the world. However, our understanding of what drives the emergence and dynamics of these variants is somewhat lacking. Most VOs have displayed minor “blips”, i.e., they increased in frequency rapidly in one location, yet this increase in frequency was also followed by a rapid decay. Only two VOCs - Alpha and Delta, displayed a more dramatic global pattern, increasing dramatically in frequency over most of the globe.

What makes these two variants unique? We first focus on non-synonymous substitutions associated with evasion from antibodies (Abs), which we denote as Abs-evasion substitutions. The Alpha variant stands out for its absence of such substitutions (with the exception of one amino-acid deletion at the NTD), which are abundant in all other VOs (Fig. 2C). This is line with the timeline during which Alpha began to spread, around the fall of 2020: the global population was not vaccinated, and it is not likely that recovered individuals constituted a large fraction of the global population. Thus, Abs-evasion substitutions bore no selective advantage in infections of naive individuals. On the other hand, we suggest that in other VOs spreading in 2020, Abs-evasion substitutions may have even incurred a fitness cost during infection of naive individuals, and this led to the limited spread of most VOs, as for example occurred with the Beta variant in Israel [28]. In contrast to Alpha, the spread of the Delta VOC is currently occurring at a very different landscape, with an increase in immunized and recovered individuals, and thus Abs-evasion substitutions may confer a significant advantage. Accordingly, results herein and by others show more infections occurring in immunized individuals as compared to what was previously observed for the Alpha variant. At this point it should be noted that current reports from Israel and elsewhere suggest that the recent increase in observed breakthrough cases with the Delta variant are likely a combination of waning immunity [29] in addition to the ability of this variant to overcome the immune response to some extent [26]. This waning may have led to somewhat reduced vaccine effectiveness against infection [30], yet effectiveness against hospitalization and severe disease remain high [31].

We now discuss the enigmatic process of how VOs are created. When the unique long branch (representing a large accumulation of substitutions) of the Alpha VOC was first noted, it was suggested that Alpha arose in an immunocompromised individual chronically infected with SARS-CoV-2 [32][33]. It has been previously observed that during treatment of such individuals with convalescent plasma or with monoclonal antibodies, rapid evolution is observed, including the accumulation of various Spike amino-acid replacements that are prevalent in VOs [32]. Accordingly, one hypothesis that emerges is that all VOs were first created in immunocompromised individual infected with SARS-CoV-2, albeit this hypothesis is very difficult to confirm, since it is impossible to detect a “patient” zero from which VOs emerged.

The Delta variant, however, appears to be quite different in this context from other VOs, especially in comparison to Alpha. The tree topology of Delta is highly structured, suggesting that its spread was a slower and more step-wise process. The Delta clade includes five newly characterized clades, suggesting two possibilities: either Delta arose early on during the pandemic, and rounds of random genetic drift led to its separation into several clades, or - selection led to the formation of these clades. The high proportion of non-synonymous substitutions during the emergence of some, but not all of these clades supports selection, yet this is inconclusive. We further note that of the five clades A-E, Delta D seems to be repeatedly gaining dominance in various countries across the globe. We go on to discuss two hypotheses: the first is that the rise in frequency of Delta D is due to founder effects, and the second is that Delta D arose due to positive selection.

One possibility that arises, is that the increase in Delta D is a combination of founder effects together with a shift in the landscape of immunized and recovered individuals across the globe. Accordingly, Delta D in itself has no selective advantage over other Delta clades; yet all Delta clades bear a selective advantage over the Alpha variant and other variants that predated Delta. In particular, it is possible that the advantage Delta bears is in its ability to overcome some of the defenses of immunized individuals as compared to Alpha, as evident from the data herein and elsewhere [34][35][36]. Thus, Delta D first increased in frequency over the period of March to May 2021 in India merely due to genetic drift, and later as the proportion of immunized/recovered individuals increased, the already prevalent Delta D took over. The caveat in this hypothesis is that infections from all Delta clades are evident already in April across the globe. It is possible that these are a biased sample from incoming travelers who did not go on to create transmission chains.

A second hypothesis is that Delta D may be under positive selection. In line with this, 82% of the lineage specific mutations of this clade are non-synonymous, similar to the proportion that characterizes VOs (Table 1, Fig. 2A). However, this clade lacks additional substitutions in S, and is characterized by seven amino-acid replacements in the ORF1a/b polyprotein. This is particularly perplexing as the lineage defining mutations of the main Delta lineage are completely devoid of mutations in ORF1a/b (Table 1). An additional non-synonymous substitution is evident in the ORF7b gene (T40I) and in the N gene (G215C), quite proximal to the 203-205 region discussed above. Notably, four of the eleven substitutions unique to the Delta D clade are observed in other VOs (Table 1), suggesting that selection may have driven the emergence of this clade. If so, we suggest that the first mutations that were fixed in the basal Delta lineage, which are mostly in the S and N genes, bore the highest selective advantage. We suggest that the additional mutations fixed in Delta D (and possibly in other clades), bore a smaller selective advantage, and were hence contingent on a larger viral population size, which was enabled due to the S and N gene mutations. This mode of step-wise evolution, from large effect to small effect mutations, is in line with evolutionary theory and has been previously observed in other RNA viruses [37], and it remains to be confirmed whether and how this pattern will be recapitulated in the future.

To summarize, we have used a comparative approach to detect a unique mode of evolution present in Delta. This step-wise mode of evolution characterized both the formation of the Delta D clade, and its subsequent spread, and is in stark contrast to the evolution observed in other VOs. In particular, the global increase in Delta frequency has occurred concurrently with the increase in Delta D, suggesting that what is now labelled as “Delta” worldwide is actually specifically the Delta D clade.

## Methods

### Ethics statement

The study was conducted according to the guidelines of the Declaration of Helsinki, and approved by the Institutional Review Board of the Sheba Medical Center institutional review board (7045-20-SMC). For the Israel cohort, patient consent was waived as the study used remains of clinical samples and the analysis used anonymous clinical data.

### Study cohorts

Global sample sequences used in this study were downloaded from GISAID (global initiative on sharing all influenza data) [38]. Sequence and patient data for the Israel Delta cohort was obtained via the Israel Consortium of SARS-CoV-2 sequencing, a national surveillance system to identify circulating and imported variants via sequencing, established in December 2020 by Israel Ministry of Health and Public Health Services. Details on sequencing and bioinformatics are given below.

### Construction of phylogenetic trees

Phylogenetic trees were constructed using NextStrain’s Augur pipeline [18]. Sequences were aligned to SARS-CoV-2 reference genome (NC_045512.2) using MAFFT [39], and a time-resolved phylogenetic tree was constructed with IQ-Tree [40] and TreeTime [41] under the generalized time reversible (GTR) substitution model and visualized with auspice [18]. Lineage nomenclature was attained from Pangolin Lineages [24]. Dating of internal nodes are reported based on NextStrain, which in turns relies on IQ-Tree [40] ancestral sequence reconstruction, dating, and assignment of confidence intervals for these dates.

### Comparison of lineage defining mutations for VOs and non-VOs

Lineage defining mutations of VOs were extracted from CoVariants [15] and outbreak.info [42]. non-VO lineages were selected from Pango lineages [43][44] based on the following criteria: at least 1,000 sequences were available, and earliest data of detection varied between March and November 2020. Of these, twelve lineages were randomly chosen with an emphasis on lineages prevalent across different continents. All lineages were derived from the B.1 lineage. For each lineage, we randomly sampled at least 250 sequences and focused on substitutions present in more than 90% of the sequences, yet absent from the lineage defining mutations of the ancestral lineages. Substitutions were then classified as extragenic, synonymous, NS, or deletions based on data from ViruSurf [45]. When estimating the fraction of NS substitutions, we grouped deletions and NS, and calculated their fraction out of both NS and synonymous. A one-sided t-test with unequal variances was used to assess the statistical significance of the higher fraction of NS in VOs.

### Characterization of Delta sub-clades

When observing the phylogenetic tree of the Delta variant in NextStrain’s global analysis (www.nextstrain.org, July 24^th^ 2021), we observed a strong separation into five distinct clades, which was recapitulated in both global and continent based builds. The five clades were all based on internal branches with at least three substitutions. Since NextStrain is based on a sample of sequences, we next verified the veracity of the clades by downloading ∼3000 sequences identified as B.1.617.2 between March 1 - July 15 2021, which were collected from Virusurf [45], without “N”s (undetermined nucleotide), to facilitate identification of *bona fide* substitutions. A representative sample from around the globe was further analyzed to avoid biases towards countries who sequence more intensely. Substitutions, as compared to the SARS-CoV-2 reference NC_045512.2, were identified in each sequence using Coronapp [46], and clustered based on similar unique substitutions that are not part of the B.1.617.2 lineage defining mutations. A similar search using ViruSurf [45] was conducted for additional clade signature substitutions, to validate each clade.

### Library preparation, sequencing and processing

Israel cohort samples were processed as follows: RNA was extracted from 200µl respiratory samples with the MagNA PURE 96 (Roche, Germany), according to the manufacturer instructions. Libraries were prepared using COVID-seq library preparation kit, as per manufacturer’s instructions (Illumina). Library validation and mean fragment size was determined by TapeStation 4200 via DNA HS D1000 kit (Agilent). Libraries were pooled, denatured and diluted to 10pM and sequenced on NovaSeq (Illumina). Fastq files were subjected to quality control using FastQC (www.bioinformatics.babraham.ac.uk/projects/fastqc/) and MultiQC [47] and low-quality sequences were filtered using trimmomatic [48]. Sequences were mapped to the SARS-CoV-2 reference genome (NC_045512.2) with Burrows-Wheeler aligner (BWA) mem [49]. Resulting BAM files were sorted and indexed using SAMtools suite [50]. Breadth and depth of sequencing was calculated from sorted BAM files using a custom python script. Consensus Fasta sequences were assembled for each sample using iVar (https://andersen-lab.github.io/ivar/html/index.html), with positions <5 nucleotides determined as Ns. Multiple alignment of sample sequences with the reference Wuhan sequence (NC_045512.2) was performed with MAFFT using default parameters [39]. All data generated via the Israel Consortium of SARS-CoV-2 sequencing, including the Israel cohort data in this manuscript, is regularly deposited and available in GISAID.

## Data Availability

All data generated via the Israel Consortium of SARS-CoV-2 sequencing, including the Israel cohort data in this manuscript, is regularly deposited and available in GISAID.

