## Supplementary figures for "The unique evolutionary dynamics of the SARS-CoV-2 Delta variant"

| | Sequence at loci 28881-28887 | Amino acid replacements in N |
|---|---|---|
| Reference sequence isolate Wuhan-Hu-1, GenBank ID MN908947 | GGGGAAC | NA |
| Clade 20B (leading to Alpha, Gamma, Lambda) | AACGAAC | R203K, G204R |
| Clade 20A (leading to Delta) | TGGGAAC | R203M |
| Clade 20C mutation (leading to Beta, Epsilon) | GGGGAAT | T205I |

**Figure S1. Substitutions in the region corresponding to amino-acids  203-205 in the N gene, across different clades.** Shown are nucleotides substitutions that occurred independently in each lineage. The three substitutions in clade 20B are thought to lead to the creation of a new transcriptional regulatory sequence (TRS) AACGAAC, whereas the other substitutions are not expected to create one.

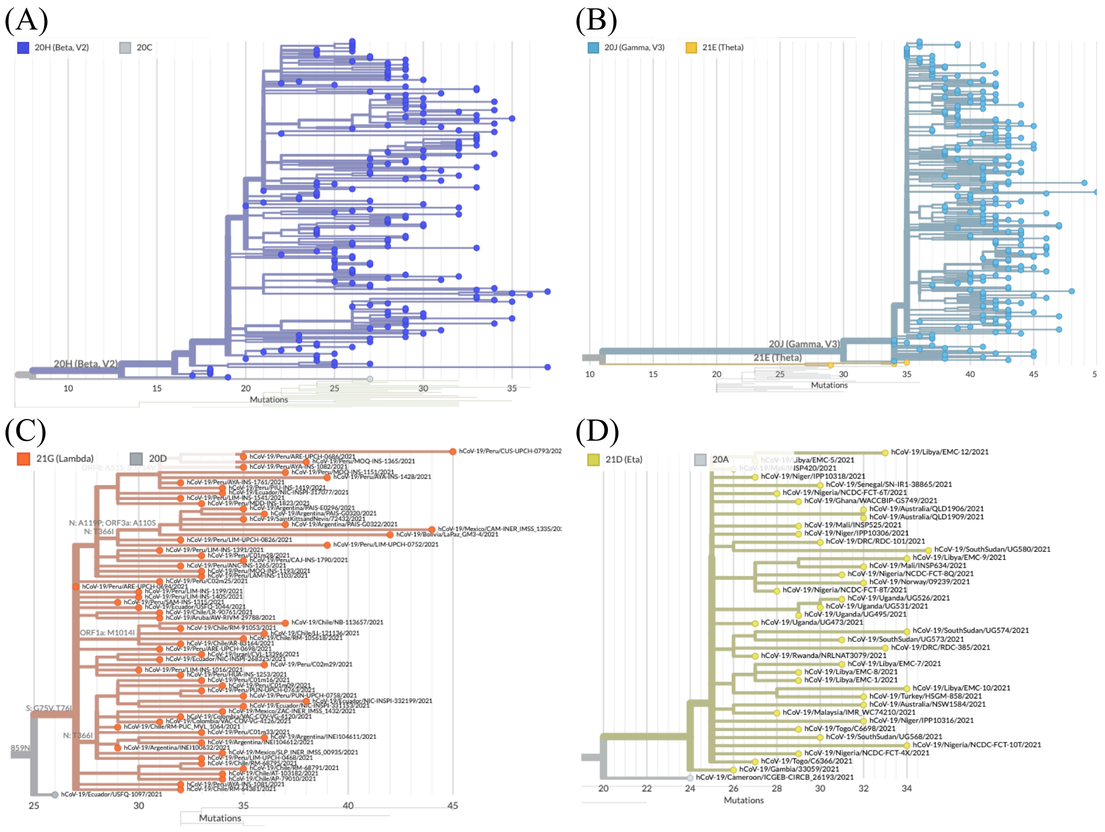


**Figure S2. Divergence-based phylogenies of the (A) Beta, (B) Gamma, (C) Lambda, (D) Eta variants.** All phylogenies show “star-like” phylogenies. Figures were generated by NextStrain on August 1 2021.

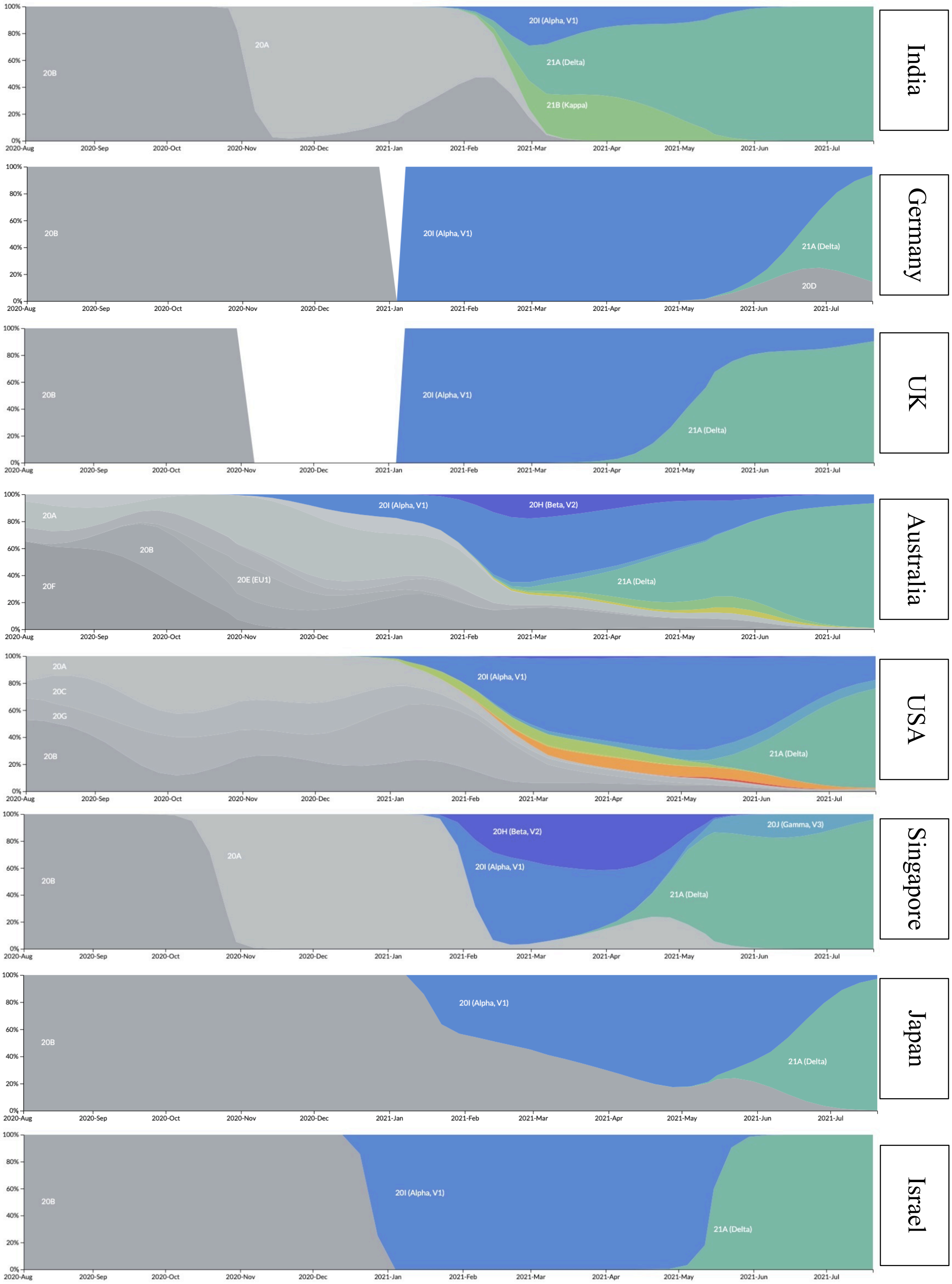


**Figure S3. Prevalence of NextStrain clades in different countries in the world, corresponding to Figure 4 of the main text.** Figures were generated by NextStrain on August 1 2021.
